# Aerobic exercise improves hippocampal blood flow for hypertensive *APOE4* carriers

**DOI:** 10.1101/2020.09.23.20199042

**Authors:** Carolyn S. Kaufman, Robyn A. Honea, Joseph Pleen, Rebecca J. Lepping, Amber Watts, Jill K Morris, Sandra A. Billinger, Jeffrey M. Burns, Eric D. Vidoni

## Abstract

**Background:** Evidence increasingly suggests cerebrovascular dysfunction plays an early and important role in the pathogenesis of Alzheimer’s disease (AD). Studies have shown the strongest known genetic risk factor for sporadic AD, *Apolipoprotein E4* (*APOE4*), may act synergistically with vascular risk factors to promote dementia development. Aerobic exercise may attenuate cognitive decline at least partially through improvements in cerebral blood flow. Therefore, exercise interventions that improve vascular health may be particularly beneficial for *APOE4* carriers.

**Objectives:** To test the hypothesis that exercise would more effectively increase hippocampal blood flow (HBF) in hypertensive *APOE4* carriers than non-carriers, we performed an analysis of arterial spin labeling MRI data (ASL-MRI) from a randomized controlled trial (secondary outcome). Additionally, we tested the hypothesis that changes in systolic blood pressure (ΔSBP) would be more negatively associated with ΔHBF for *APOE4* carriers than non-carriers.

**Methods:** We assigned cognitively normal adults (65–87 years) to a 52-week aerobic exercise intervention or education only. Genotyping was performed by Taqman SNP allelic discrimination assay. ASL-MRI measured HBF before and after the 52-week intervention. We selected participants with hypertension at enrollment (n = 44), defined as SBP ≥ 130mmHg or diastolic blood pressure (DBP) ≥ 80mmHg.

**Results:** A two-way ANCOVA showed a significant interaction between *APOE4* carrier status and treatment group on change in HBF (ΔHBF) over the 52 weeks, controlling for age and sex (*p* = 0.040). For *APOE4* carriers, ΔHBF was significantly (*p* = 0.006) higher for participants who underwent the exercise intervention (4.09 mL/100g/min) than for the control group (−2.08 mL/100g/min). There was no difference in ΔHBF between the control (−0.32 mL/100g/min) and exercise (−0.54 mL/100g/min) intervention groups for *APOE4* non-carriers (p = 0.918). Additionally, a multiple linear regression showed a significant interaction between ΔSBP and *APOE4* carrier status on ΔHBF (*p* = 0.035), with a reduction in SBP associated with an increase in HBF for *APOE4* carriers only.

**Conclusions:** Aerobic exercise significantly improved HBF for hypertensive *APOE4* carriers only. Additionally, only *APOE4* carriers exhibited an inverse relationship between ΔSBP and ΔHBF. This suggests exercise interventions, particularly those that lower SBP, may be beneficial for individuals at highest genetic risk of AD.

## INTRODUCTION

Evidence increasingly points to an early, primary role of cerebrovascular dysfunction in the pathogenesis of late-onset Alzheimer’s disease (AD) [1-3].Considering the well-established benefits of aerobic exercise on vascular health [4], exercise may act mechanistically through improvements in vascular function to reduce dementia risk. Indeed, intervention trials have shown exercise may improve cognitive function at least partially through increasing cerebral blood flow (CBF) [5-8], particularly in the hippocampus (HBF) [9-11].

The strongest known genetic risk factor for AD, the *APOE4* allele, may act synergistically with poor vascular health to increase dementia risk [12-18]. This suggests interventions that improve systemic vascular health may be particularly beneficial for *APOE4* carriers [19]. Some studies have shown *APOE4* carriers benefit more from exercise [20-23], but others have suggested less improvement in *APOE4* carriers [24, 25], meriting further exploration.

We performed an analysis of a secondary outcome from a randomized controlled trial in which cognitively normal older adults were assigned to a 52-week aerobic exercise intervention or to education only. The primary outcome of the trial was the effect of a 52-week aerobic exercise program on AD pathophysiology (β-amyloid burden) [26]. For the present analysis, we selected only participants with hypertensive blood pressure at the time of enrollment, defined as systolic blood pressure (SBP) ≥ 130 mmHg or diastolic blood pressure (DBP) ≥ 80 mmHg [27]. We hypothesized that 1) the exercise intervention would be more effective in increasing HBF in *APOE4* carriers than non-carriers, and 2) reductions in SBP would have a greater impact on improving HBF for *APOE4* carriers than for non-carriers.

## METHODS

### Study design

The Alzheimer’s Prevention through Exercise study (APEx) was a 52-week clinical trial of aerobic exercise in cognitively normal older adults. The primary outcome was change in β-amyloid deposition, and these results have been published previously [26]. The present study is an analysis of the secondary outcome of regional arterial spin labeling (ASL) MRI data, specifically HBF, for the participants in the APEx clinical trial (NCT02000583).

### Participants

Participants were required to meet the following inclusion criteria: 65 years and older, sedentary or underactive as defined by the Telephone Assessment of Physical Activity [28], on stable medications for at least 30 days, willingness to undergo an 18F-AV45 PET scan for cerebral β-amyloid load and learn the result (elevated or non-elevated), willingness to perform prescribed exercise (or not) for 52 weeks at a community fitness center, and ability to complete graded maximal exercise testing with a respiratory exchange ratio >=1.0. Exclusion criteria included insulin-dependence,significant hearing or vision problems, clinically evident stroke, cancer in the previous 5 years (except for localized skin or cervical carcinomas or prostate cancer), change in blood pressure medication within the last 30 days, or recent history (<2 years) of major cardiorespiratory, musculoskeletal or neuropsychiatric impairment. During in-person screening, a clinician of the University of Kansas Alzheimer’s Disease Center performed a clinical assessment that included a Clinical Dementia Rating, the Uniform Data Set neuropsychiatric battery, and other tests [29, 30].

### Neuroimaging assessments

Individuals who consented to screening underwent florbetapir 18F-AV45 (370 MBq) PET scans. β-amyloid status was disclosed to all participants [31]. We enrolled those participants with cortical-to-cerebellar β-amyloid burden greater than 1 because these participants may have accelerated β-amyloid deposition and memory decline [32]. At baseline, enrollees had a T1-weighted MRI of the brain (Tesla Skyra scanner; MP-RAGE 1×1×1.2mm voxels, TR = 2300ms, TE = 2.98ms, TI = 900ms, FOV 256×256mm, 9° flip angle; ASL single-shot EPI 3.8×3.8×4.0mm,TR = 3400ms, TE = 13ms, TI = 700ms, FOV 240×240mm, 90° flip angle) with regional volumes parcellated and extracted using the CAT12 toolbox (http://www.neuro.uni-jena.de/cat/) and the Automatic Anatomic Labeling atlas. ASL-MRI data were processed using the ASLTbx for SPM12 [33]. All neuroimaging assessments were repeated at 52 weeks.

### Physiological assessments

At the baseline study visit, the participant sat at rest for 5 minutes before BP was measured twice with one minute of rest between measurements (Axia TRIA Touch Screen Patient Monitor, Association for the Advancement of Medical Instrumentation/American National Standards Institute performance standards SP10:2002). We averaged the two resting SBP and two resting DBP to determine one average baseline SBP/DBP, which was used for grouping into hypertensive and normotensive categories based on the most recent guidelines published by the American College of Cardiology and American Heart Association (ACC/AHA) [27].Before beginning the study, participants performed graded maximal exercise testing on a treadmill to maximal capacity or volitional termination to quantify cardiorespiratory fitness (VO_2_max) [34]. The BP measurement and graded maximal exercise test were repeated at 52 weeks.

### Cognitive assessments

A trained psychometrist performed a comprehensive cognitive test battery at baseline and 52 weeks, employing validated, alternate versions of tests every other visit. We created composite scores for three cognitive domains (executive function, verbal memory, visuospatial processing) using Confirmatory Factor Analysis. Scores were standardized to baseline so subsequent scores could be interpreted relative to baseline. The executive function composite score was made up of verbal fluency [35], Trailmaking Test B [36], Digit Symbol Substitution test [37], and the interference portion of the Stroop test [38]. The verbal memory composite score was made up of the Logical Memory Test immediate and delayed [37], and the Selective Reminding Test [39]. The visuospatial composite score was made up of scores from Block Design [37], space relations, the paper folding test, hidden pictures, and identical pictures [40]. Missing data in the factor analysis were accounted for using full information maximum likelihood algorithm.

### Intervention

Participants were randomized in a 2:1 ratio to either 150 minutes per week of supported moderate intensity aerobic exercise or standard of care education. The education control group was provided standard exercise public health information but was otherwise not supported nor prohibited from exercise. For those randomized to the aerobic exercise group, the intervention was conducted at their nearest study-certified exercise facility with the support of certified personal trainers. The intervention group was asked to refrain from changing their regular physical activities other than those prescribed by the study team. Participants exercised 3–5 days a week at an intensity that began at 40 – 55% of Heart Rate Reserve (% of the difference between maximal and resting) and was increased by 10% every 3 months.

### APOE genotype determination

Whole blood was collected and stored at -80C until genetic analyses could be conducted. To determine *APOE* genotype, frozen whole blood was assessed using a Taqman single nucleotide polymorphism (SNP) allelic discrimination assay (ThermoFisher). *APOE4, APOE3*, and *APOE2* alleles were distinguished using Taqman probes to the two *APOE*-defining SNPs, rs429358 (C_3084793_20) and rs7412 (C_904973_10). The term “*APOE4* carrier” was used to describe the presence of 1 or 2 *APOE4* alleles. Since *APOE2* is associated with reduced AD risk, all *APOE2* carriers were excluded from the analysis (whether homozygous or paired with a different *APOE* allele).

### Standard protocol approvals, registrations, and patient consents

All study (ClinicalTrials.gov, NCT02000583; trial active between 11/1/2013– 11/6/2019) procedures were approved by the KU Institutional Review Board and complied with the Declaration of Helsinki. Written informed consent was obtained from all participants.

### Statistical approach

All statistical analyses were performed using SPSS Statistics (IBM). Baseline group differences were assessed by independent t-test, chi-square test for homogeneity, Mann Whitney U, Kruskal-Wallis H test, or one-way ANOVA, as appropriate. A multiple linear regression further characterized the relationship between SBP, DBP and baseline HBF. A two-way ANCOVA was conducted to examine the effects of *APOE4* carrier status and Treatment Group (exercise or control) on ΔHBF, after controlling for age and sex. The significant interaction term was followed up by an analysis of simple main effects using a Bonferroni adjustment (p < 0.025). A multiple regression was run to predict ΔHBF from sex, *APOE4* carrier status, ΔSBP, and the interaction between ΔSBP and *APOE4* carrier status. Age, ΔDBP and the interaction term for ΔDBP and *APOE4* carrier status worsened the predictive value and were therefore excluded from the model. Two-way ANCOVAs were utilized to examine the effects of *APOE4* carrier status and Treatment Group (exercise or control) on change in cognitive functioning (visuospatial, executive and memory), ΔSBP, ΔDBP, change in hippocampal volume, and change in VO2max (ΔVO2mx), after controlling for age and sex. A Pearson’s Product-Moment Correlation assessed the relationship between ΔHBF and change in verbal memory function (ΔVM) for the *APOE4* carriers who underwent the exercise intervention.

## RESULTS

A total of 109 participants (93%: control n = 34, aerobic exercise n = 75) completed the study (Figure 1). Genotyping was not completed for 3 participants. *APOE2* carriers (n = 14) and participants with incomplete ASL MRI data (n = 4) were excluded from the present analysis. Of the remaining 88 participants, 44 had hypertension at the baseline visit, classified due to having SBP ≥ 130 mmHg (n = 22), DBP ≥ 80 mmHg (n = 5), or both (n = 17) [27]. Sixteen participants completed less than 80% of the prescribed exercise, but all participants were included in the analyses regardless of the percent exercise intervention prescription completed.

**Figure 1.**
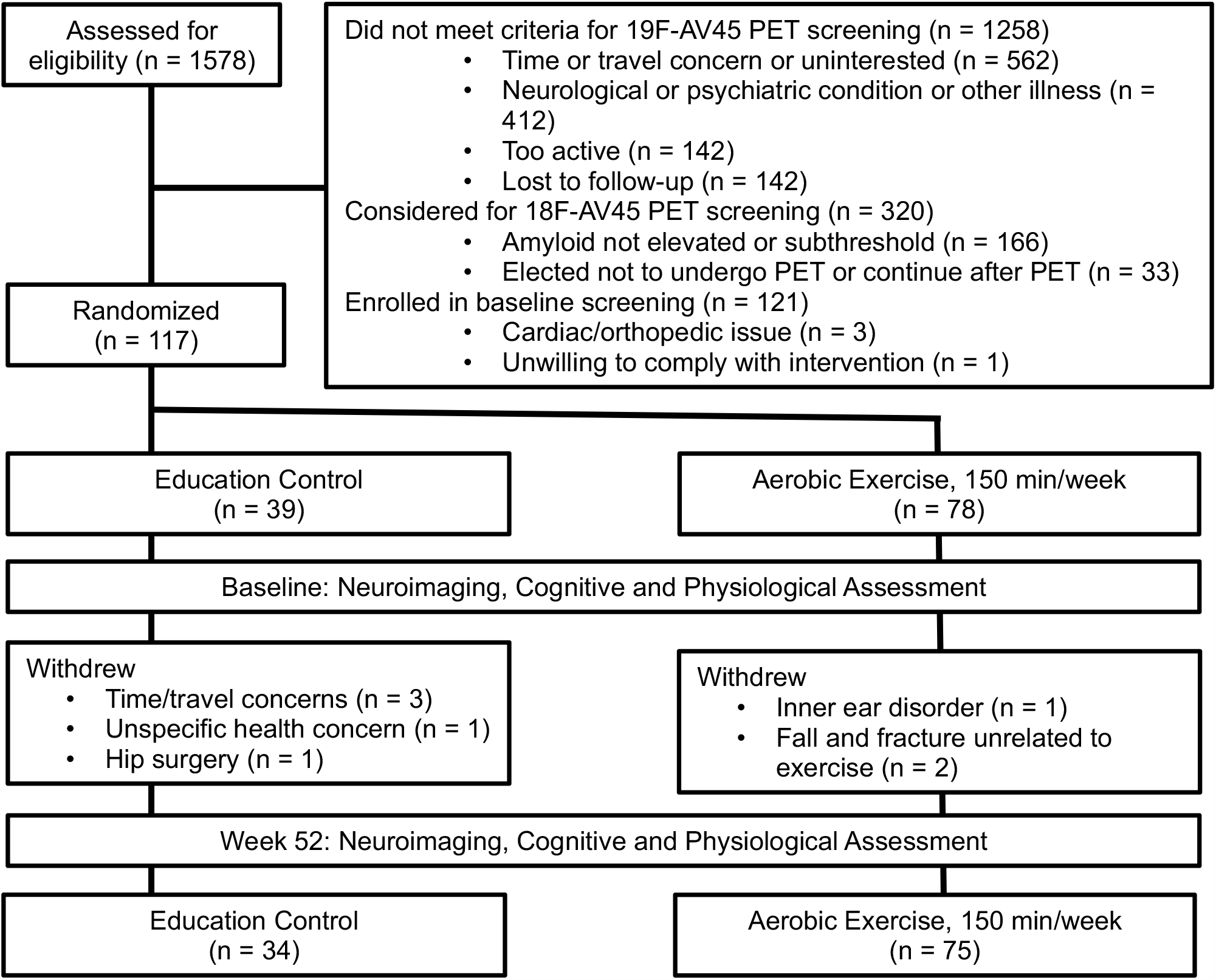
Flowchart of Alzheimer’s Prevention through Exercise (APEx) study participants

### Baseline characteristics of hypertensive and normotensive participants

At baseline, HBF was significantly lower in the hypertensive participants (30.17 ±6.06 mL/100g/min) than the normotensive participants (34.00 ± 9.54 mL/100g/min) (*p* = 0.028). A multiple linear regression showed this difference was driven by baseline SBP. Specifically, the regression analysis, controlling for age, sex and *APOE4* carrier status, significantly predicted HBF (*F*(5,82) = 2.657, *p* = 0.028) with SBP contributing significantly to the model (*B* = -0.157, *p* = 0.035) and DBP having a non-significant effect (*B* = 0.086, *p* = 0.461). There were no significant differences between the hypertensive and normotensive participants in age, sex, education, VO_2_max or hippocampal volume (Table 1).

**Table 1.**
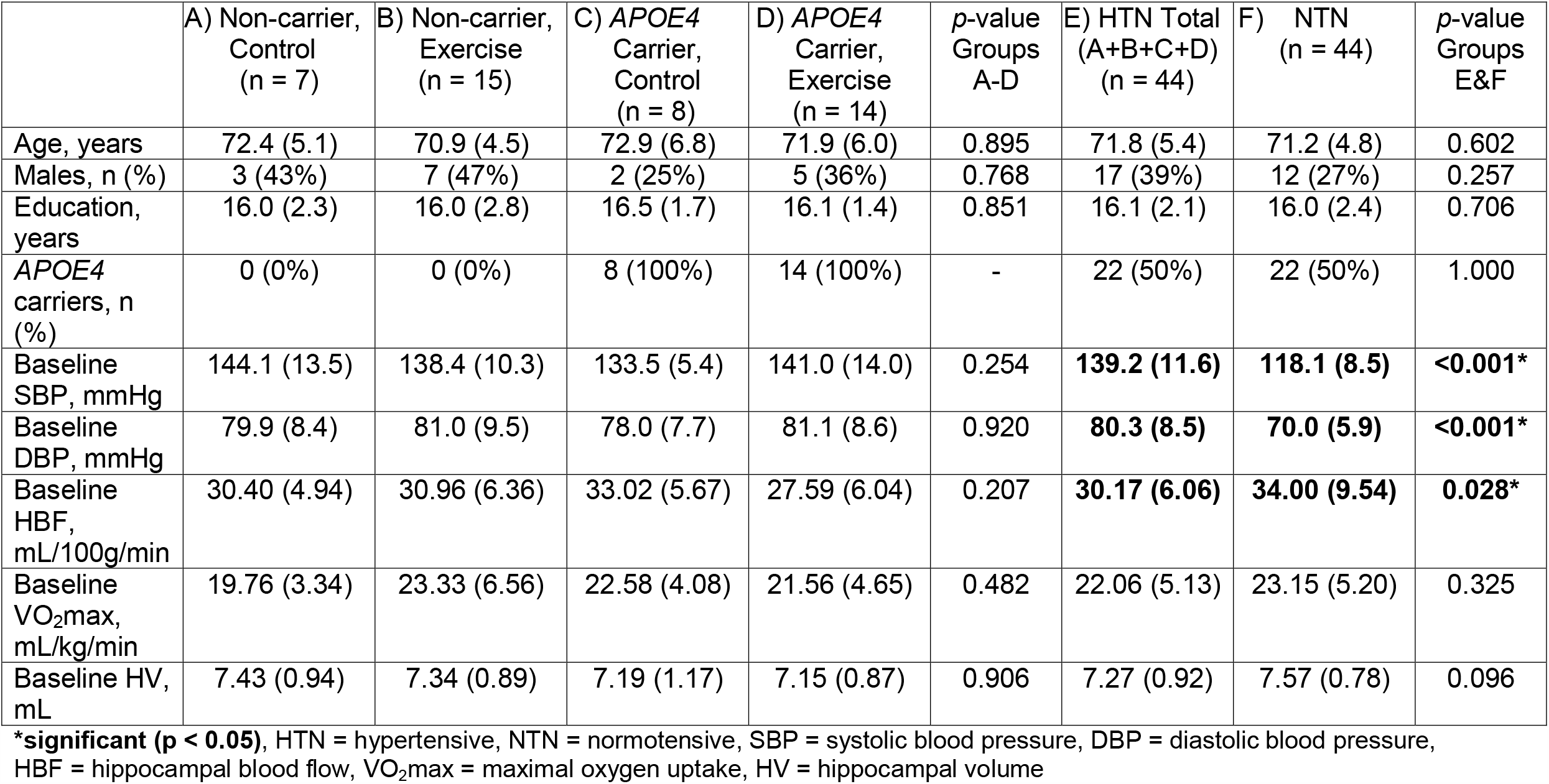
Baseline characteristics by group

### Baseline characteristics by intervention and APOE4 carrier status

Only the participants with hypertension at the baseline visit (n = 44) were included in further analyses. Half of these participants were *APOE4* carriers (*APOE3/APOE4*, n = 20; *APOE4/APOE4*, n = 2). At baseline, there were no significant differences among *APOE4* non-carriers assigned to the control group (n = 7), *APOE4* non-carriers assigned to exercise (n = 15), *APOE4* carriers assigned to the control group (n = 8), and *APOE4* carriers assigned to exercise (n = 14) in age, sex, education, SBP, DBP, HBF, maximal oxygen uptake (VO_2_max), or hippocampal volume (Table 1).

### ΔHBF with exercise intervention

The two-way ANCOVA met the assumptions of homoscedasticity and homogeneity. There were no outliers in the data, as assessed by no cases with studentized residuals greater than ±3 SD. Studentized residuals were normally distributed, as assessed by Shapiro-Wilk’s test (*p* > .05). Means, adjusted means (for age and sex), SD and standard errors are presented in Table 2 and shown graphically in Figure 2. There was a significant two-way interaction between *APOE4* carrier status and Treatment Group on ΔHBF, while controlling for age and sex, *F*(1, 38) = 4.504, *p* =0.040, partial η^2^ = 0.106. Therefore, an analysis of simple main effects for *APOE4* carrier status and Treatment Group was performed with statistical significance receiving a Bonferroni adjustment and being accepted at the *p* < 0.025 level.

**Table 2.** Mean change in HBF (ΔHBF) from baseline to 52 weeks

| ΔHBF<br>(mL/100 g/min) | Non-carrier<br>Control<br>(n = 7) | Non-carrier<br>Exercise<br>(n = 15) | *APOE4* Carrier<br>Control<br>(n = 8) | *APOE4* Carrier<br>Exercise<br>(n = 14) |
| --- | --- | --- | --- | --- |
| Mean | -0.19 | -0.48 | -2.22 | +4.05 |
| (SD) | 2.14 | 4.06 | 4.32 | 6.21 |
| Mean<sub>adjusted</sub> | -0.32 | -0.54<sup>*</sup> | -2.08<sup>+</sup> | +4.09<sup>*,+</sup> |
| (SE) | 1.79 | 1.23 | 1.69 | 1.26 |

SD = Standard Deviation; SE = Standard Error; <sup>\*</sup> $p = 0.013$ ; <sup>+</sup> $p = 0.006$
Mean<sub>adjusted</sub> represents the group means, adjusted for age and sex, for participants divided by *APOE4* carrier status and clinical trial arm (exercise intervention or control). Significant results of the simple main effects analysis that followed up the significant two-way ANCOVA interaction are indicated by superscripts (<sup>\*</sup> and <sup>+</sup>). All other pairwise comparisons were not significant.

**Figure 2.**
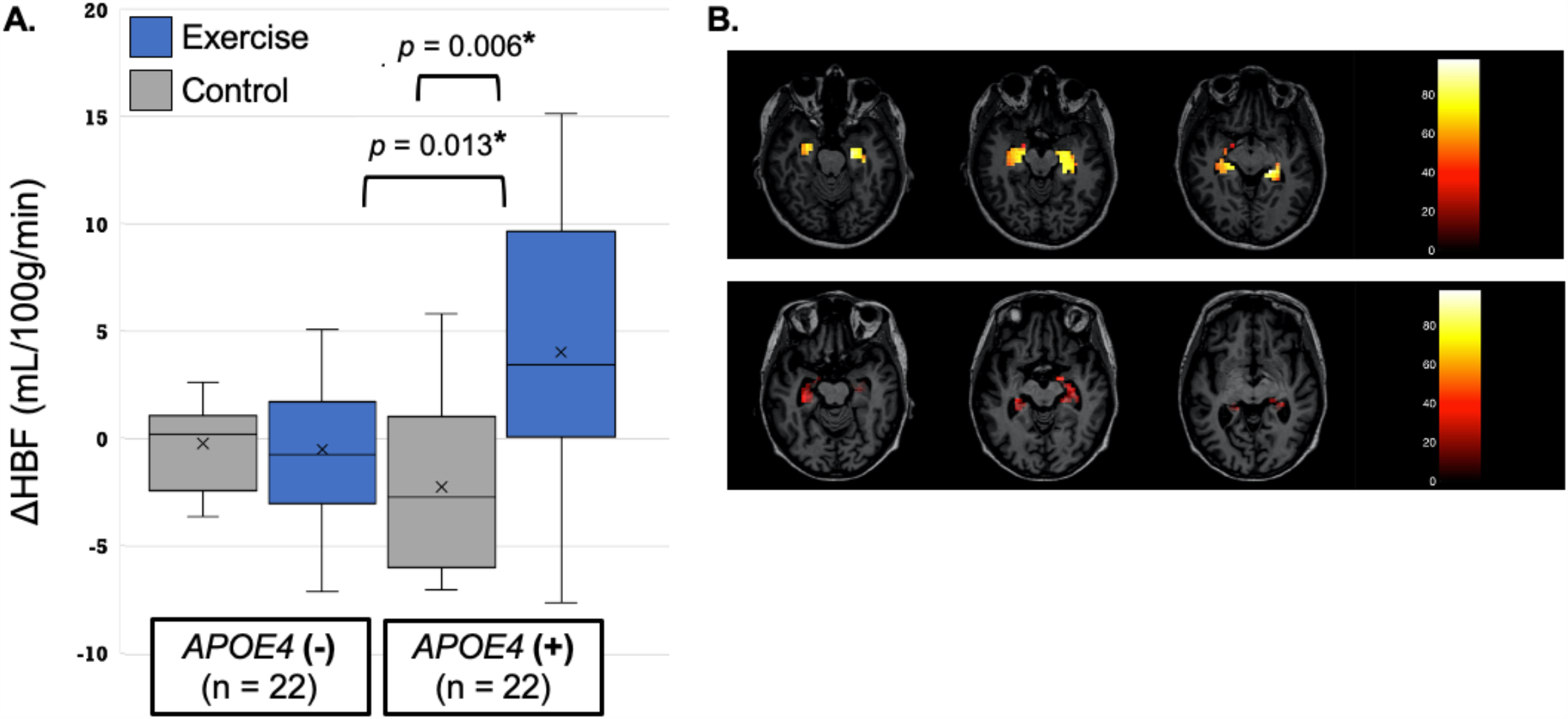
Mean change in hippocampal blood flow (ΔHBF) from baseline to 52 weeks **A)** Among *APOE4* carriers, those who underwent the exercise intervention had a significantly larger ΔHBF (increased HBF) over the 52 weeks than the control group (*p* = 0.006). Additionally, within the exercise intervention arm, ΔHBF was significantly larger for the *APOE4* carriers than the non-carriers (*p* = 0.013). There were no other significant differences between groups. X = mean; horizontal line = median **b)** Representative arterial spin labeling MRI (ASL-MRI) scans from a participant with high (top panel) and a participant with low (bottom panel) HBF.

The effect of Treatment Group on ΔHBF was significant for the *APOE4* carriers, *F*(1, 38) = 8.597, *p* = 0.006, partial η^2^ = 0.184. Specifically, for the *APOE4* carriers, adjusted mean ΔHBF was higher for participants who underwent the exercise intervention (4.09 mL/100g/min) than for the control group (−2.08 mL/100g/min), a significant difference of 6.17 (95% CI, 1.91 to 10.44) mL/100g/min. The effect of Treatment Group was not significant in the *APOE4* non-carriers, *F*(1, 38) = 0.011, *p* = 0.918, partial η^2^ < 0.0001.

The effect of *APOE4* carrier status on ΔHBF in the exercise group was significant, *F*(1, 38) = 6.853, *p* = 0.013, partial η^2^ = 0.153. Specifically, for participants who underwent the exercise intervention, adjusted mean ΔHBF was higher for the *APOE4* carriers (4.09 mL/100g/min) than the non-carriers (−0.54 mL/100g/min), a significant difference of 4.63 (95% CI, 1.05 to 8.22) mL/100g/min. In the control group, the effect of *APOE4* carrier status on ΔHBF was not significant, *F*(1, 38) = 0.514, *p* = 0.478, partial η^2^ = 0.013.

Overall, the 52-week exercise intervention improved mean HBF for the *APOE4* carriers from 27.59 to 31.64 mL/100g/min. Over this same time period, the *APOE4* carriers in the control group experienced a decline in HBF from 33.02 to 30.80 mL/100g/min, and the *APOE4* non-carriers remained relatively stable in both the control (30.40 to 30.22 mL/100g/min) and exercise intervention (30.96 to 30.48 mL/100g/min) groups. It is thus noteworthy that the *APOE4* carriers in the exercise intervention had the lowest mean HBF at baseline but the highest HBF after the intervention, although these groups differences were not significant at baseline (*p* = 0.207) or post-intervention (*p* = 0.961).

### ΔHBF with change in systolic blood pressure (ΔSBP)

We ran a multiple linear regression analysis to predict ΔHBF over the 52-week period from sex, *APOE4* carrier status, ΔSBP, and the interaction between ΔSBP and *APOE4* carrier status. The regression analysis met the assumptions of linearity, normality, homoscedasticity, and independence of residuals. There was no multicollinearity. There were no outliers, as assessed by no studentized deleted residuals greater than ±3 SD. The multiple linear regression model significantly predicted ΔHBF, *F*(4, 39) = 3.134, *p* = 0.025, R^2^ = 0.243, adjusted *R*^*2*^ = 0.166. Regression coefficients and standard errors can be found in Table 3.

**Table 3.** Regression model for 52-week change in hippocampal blood flow (ΔHBF)

| Variable | *B* | SE<sub>*B*</sub> | $\beta$ | *p*-value |
| --- | --- | --- | --- | --- |
| Intercept | -1.529 | 1.232 |  | 0.222 |
| Sex (Male) | 2.369 | 1.478 | 0.226 | 0.117 |
| ΔSBP | -0.015 | 0.065 | -0.041 | 0.815 |
| *APOE4* carrier status (+) | 2.479 | 1.458 | 0.243 | 0.097 |
| ΔSBP x *APOE4* carrier status (+) | -0.243 | 0.111 | -0.376 | 0.035<sup>*</sup> |

<sup>\*</sup>significant ( $p < 0.05$ ), ΔSBP = change in systolic blood pressure
The multiple linear regression model for participants with baseline hypertension (N = 44) significantly predicted ΔHBF, $F(4, 39) = 3.134$ , $p = 0.025$ , $R^2 = 0.243$ , adjusted $R^2 = 0.166$ .

There was a significant interaction between ΔSBP and *APOE4* carrier status on ΔHBF (*p* = 0.035). The relationship between ΔSBP and ΔHBF is shown graphically for *APOE4* carriers and non-carriers in Figure 3. For *APOE4* carriers, reductions in SBP over the year of the study resulted in higher HBF, while increases in SBP (reflecting further elevation of already elevated blood pressure) resulted in decreased HBF. In contrast, there was no relationship between changes in SBP and HBF for the *APOE4* non-carriers.

**Figure 3.**
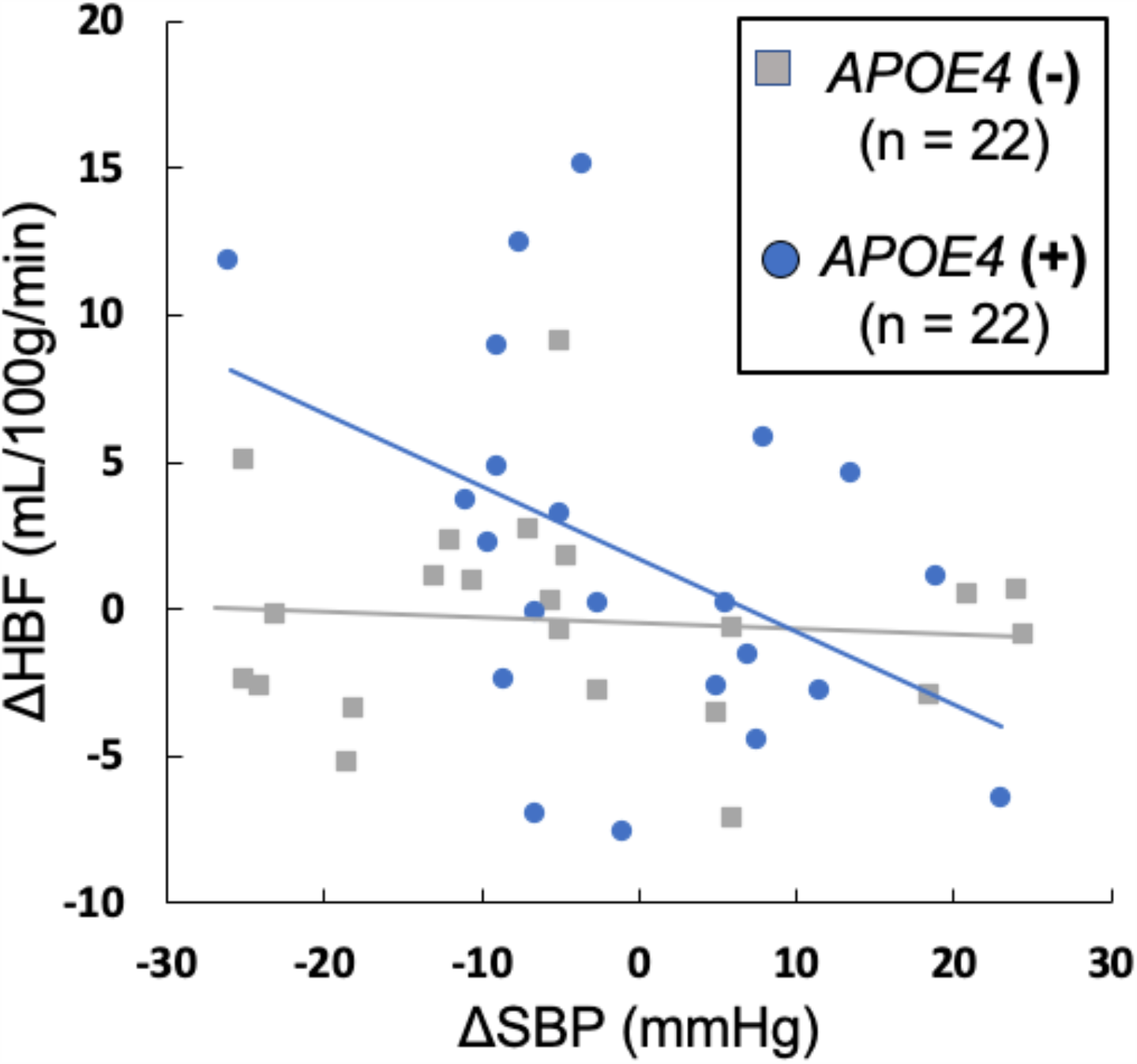
Relationship between change in systolic blood pressure (ΔSBP) and hippocampal blood flow (ΔHBF) Over the 52-week clinical trial period, ΔSBP was inversely associated with ΔHBF for the *APOE4* carriers. There was no relationship between ΔSBP and ΔHBF for the *APOE4* non-carriers. This suggests reductions in SBP in older adults with baseline hypertension may preferentially improve HBF for *APOE4* carriers.

### Cognitive function changes

There was no significant two-way interaction between *APOE4* carrier status and Treatment Group on change in visuospatial functioning (*p* = 0.755), executive functioning (*p* = 0.841), or verbal memory (ΔVM, *p* = 0.434), suggesting no group differences in mean change in cognitive scores from baseline to post-intervention.However, there was a significant positive correlation between ΔHBF and ΔVM for the *APOE4* carriers who underwent the exercise intervention (*r* = 0.561, *p* = 0.037). That is, improvements in HBF from the exercise intervention correlated with improved verbal memory performance in *APOE4* carriers.

### Other physiological changes

There was no significant two-way interaction between *APOE4* carrier and Treatment Group for ΔSBP (*p* = 0.058), ΔDBP (*p* = 0.260), or change in hippocampal volume (*p* = 0.767). There was no significant two-way interaction between *APOE4* carrier status and Treatment Group on ΔVO_2_max (*p* = 0.116). However, the main effect of Treatment Group on ΔVO_2_max was significant (*p* = 0.002). Specifically, participants who underwent the exercise intervention had a mean ΔVO_2_max of 2.729 mL/kg/min, which was significantly higher than the control group mean ΔVO_2_max of 0.407 mL/kg/min. These findings show the exercise intervention improved cardiorespiratory fitness and that this effect was not different for *APOE4* carriers and non-carriers.

## Discussion

In this analysis of a secondary outcome from a randomized controlled trial, we report that an aerobic exercise intervention selectively improved HBF for hypertensive *APOE4* carriers. Additionally, we demonstrate that reductions in SBP for hypertensive individuals were tied to improvements in HBF for *APOE4* carriers only. Finally, we found that these improvements in HBF were correlated with improved verbal memory performance.

People with AD have lower cerebral blood flow (CBF) than age-matched controls [2, 41]. A recent study involving over 7,700 scans from 1,171 people in the Alzheimer’s Disease Neuroimaging Initiative (ADNI) database found cerebrovascular dysregulation was the earliest pathological event during AD development, followed by changes in β-amyloid deposition, metabolic dysfunction, functional impairment and structural atrophy [1]. Therefore, interventions that maintain or improve CBF with aging may prevent or delay dementia development, and this may be particularly true for regions closely involved in the disease process, such as the hippocampus [42, 43]. In the present study, we demonstrate for the first time that an aerobic exercise intervention may improve HBF selectively for *APOE4* carriers. This HBF improvement could help prevent or delay AD for these individuals at highest known genetic risk, considering the growing evidence that CBF reductions precede measurable cognitive decline [1, 44] and likely contribute causally to dementia pathogenesis [3, 45-47]. Additionally, *APOE4* carriers have been shown to experience an accelerated age-related CBF decline [48, 49], and some studies suggest cerebrovascular dysfunction may act synergistically with the *APOE4* allele to promote cognitive decline [12, 50]. If true, this would mean CBF maintenance is even more important for *APOE4* carriers than non-carriers in order to prevent dementia, strengthening the clinical relevance of our current findings.

Observational studies have shown higher levels of physical activity and exercise and greater cardiorespiratory fitness are associated with reduced brain atrophy, cognitive decline and dementia risk [51-60]. One such study published earlier this year combined data from the Chicago Health and Aging Project and the Memory and Aging Project and concluded that adults should participate in at least 150 minutes of moderate-to-vigorous physical activity per week to reduce dementia risk [60], which is notably the same exercise prescription utilized in the intervention arm of our trial.Previous randomized controlled trials have shown promising results for exercise in preventing cognitive decline [8, 20, 25, 61-65], and a recent systematic review concluded exercise improves cognitive function in people 50+ years of age, independent of baseline cognitive status [66]. A better understanding of the mechanisms through which this occurs would allow tailoring and targeting of exercise interventions for populations most likely to benefit. One important mechanism may be through improvements in CBF, particularly in the hippocampus. Indeed, aerobic exercise has been shown to increase HBF both acutely [67] and chronically [7, 9, 68], with increased HBF correlating with cognitive gains [10, 11]. However, to our knowledge, we are the first to report a selective increase in HBF with exercise for *APOE4* carriers. The positive correlation between ΔHBF and ΔVM for our *APOE4* intervention group suggests these improvements in HBF may play a role in preventing cognitive decline.

There have been conflicting reports on the influence of the *APOE4* allele in exercise-induced changes in cognitive function and brain health. One randomized trial involving 170 older adults with self-reported memory problems suggested less cognitive benefit from exercise for *APOE4* carriers [25]. However, a more recent trial of 200 patients with mild AD found *APOE4* carriers experienced more improvement in cognitive function after an exercise intervention than non-carriers [20]. Observational studies have likewise produced conflicting findings. For example, one study of cognitively-normal older adults concluded high physical activity level may preserve hippocampal volume in *APOE4* carriers selectively [22]. However, a different cross-sectional study including older adults with and without AD found *APOE4* carrier status did not influence the relationship between cardiorespiratory fitness and brain atrophy [59]. Our current findings expand upon these conflicting results by providing further evidence for a preferential benefit of exercise for *APOE4* carriers, specifically in regional CBF improvement.

Hypertension may act mechanistically through cerebrovascular dysfunction to cause brain pathology [69]. Unlike family history, hypertension is a modifiable risk factor for dementia, and lowering blood pressure is thus an enticing intervention strategy to slow or prevent cognitive decline. The landmark Systolic Blood Pressure Intervention Trial Memory and Cognition in Decreased Hypertension (SPRINT-MIND) trial published in 2019 demonstrated that aggressive BP lowering (SBP < 120 mmHg) through medication significantly reduced the risk of MCI in 9,361 older adults [70]. Notably, this was the first instance of an intervention of any type effectively reducing mild cognitive impairment (MCI) incidence in a large population [70]. Although the mechanism through which aggressive SBP lowering prevented cognitive decline was beyond the scope of the SPRINT-MIND trial, other studies have suggested reducing BP in people with hypertension may act mechanistically by augmenting CBF. For example, one group found intensive BP lowering in cognitively-normal older adults significantly increased gray matter CBF [71]. More recently, the Nilvadipine in AD (NILVAD) trial reported that reducing SBP significantly increased HBF in adults with mild-to-moderate AD, suggesting SBP reductions may provide benefit by increasing blood flow to the hippocampus specifically [72]. In the current study, baseline assessment of the entire population (hypertensive and normotensive participants, n = 88) showed higher SBP was related to lower HBF, even when controlling for age, sex and *APOE4* carrier status. This finding provides further evidence for the connection between hypertension and reduced HBF in older adults.

By selecting only individuals with hypertension at baseline (defined using the ACC/AHA Clinical Practice Guidelines [27]), the present analysis included older adults at high baseline vascular risk. We found that reductions in SBP for these participants with initially elevated BP were tied to improvements in HBF for the *APOE4* carriers only. This is in line with previous literature showing elevated blood pressure acts synergistically with *APOE4* carrier status to impair cognitive function [13, 17, 73] and promote brain pathology [16, 18]. Furthermore, while the NILVAD sub-study did not assess *APOE4* carrier status [72], it seems plausible that the observed improvement in HBF from SBP reduction could have been driven by a large proportion of *APOE4* carriers in the study population, considering the high prevalence of *APOE4* in AD (∼65% of people with AD compared to ∼25% of the general population) [74]. Regardless, the existing literature combined with our current data provide strong evidence for a synergistic relationship between the *APOE4* allele and peripheral BP on promoting brain pathology and cognitive dysfunction, suggesting interventions to lower BP are particularly important for this patient population.

Our study has a number of limitations. This was an analysis of a secondary outcome of a clinical trial with different primary aims [26], and the findings should therefore be interpreted with caution. All participants were healthy and cognitively normal, which may complicate the generalizability of our data to patient populations. Although changes in verbal memory did correlate with changes in HBF over the 52-week period for the *APOE4* carriers who exercised, there were no differences among intervention groups in mean change in cognitive functioning for any domain, which means we cannot state that exercise improved cognition. The ACC/AHA guidelines recommend BP measurements taken on at least two separate occasions in order to diagnose hypertension. For the current study, we obtained two BP measurements on a single occasion. Therefore, some participants may have been included in our hypertensive group who would not have met the criteria if given a second reading on a separate occasion. Finally, although our trial included a longer intervention than the majority of previously-published exercise intervention trials [5, 8, 10, 20, 25, 61, 63, 64, 75], the follow-up period of 52 weeks may still be too short to sufficiently characterize long-term clinical relevance of the exercise intervention. Future studies that follow participants for many years post-intervention would be better suited to assess clinical implications such as dementia risk.

In the current study, we report an aerobic exercise intervention improved HBF for cognitively normal hypertensive *APOE4* carriers (but not non-carriers) and that ΔHBF over the 52-week intervention period was positively correlated with ΔVM performance for this group. Additionally, we show ΔSBP was inversely associated with ΔHBF for *APOE4* carriers only. These findings suggest aerobic exercise interventions, especially those that lower SBP, may be particularly beneficial for *APOE4* carriers with baseline hypertension. This knowledge could inform the design and execution of future interventional trials.

## Data Availability

This trial was prospectively registered with ClinicalTrials.gov (NCT02000583). Anonymized data will be shared by request from any qualified investigator.

